# Continuous neural control of a 2-DOF ankle-foot prosthesis enables dynamic obstacle maneuvers after transtibial amputation

**DOI:** 10.1101/2025.11.25.25340897

**Authors:** Tsung-Han Hsieh, Hyungeun Song, Christopher Shallal, Daniel V. Levine, Seong Ho Yeon, Junqing Qiao, Tony Shu, Matthew J. Carty, John McCullough, Hugh M. Herr

## Abstract

Bionic reconstruction techniques that employ surgical neuroprosthetic interfaces, biomimetic control systems, and powered mechatronics have enabled versatile and biomimetic legged gait without reliance on intrinsic gait controllers. However, relative emphasis has been placed on the emulation of sagittal plane biomechanics while neglecting to provide control over frontal plane mechanics critical for terrain adaptation. Here, we present a 2-degree-of-freedom (DOF) bionic reconstruction at the transtibial amputation level that enables continuous neural control of both sagittal and frontal ankle and subtalar joint mechanics. To demonstrate its capabilities in a case study design, we integrated a 2-DOF robotic ankle-foot device via surface electromyographic electrodes to an individual provisioned with a surgical neuroprosthetic interface that augments residual muscle afferents. The subject was able to neurally control both degrees of freedom to regain nominal gait mechanics during both level-ground walking and continuous cross-slope navigation. Furthermore, the subject strategically traversed an obstacle course by dynamically hopping between a series of discrete cross-slope blocks, adapting to the slopes, and responding to rapid foot slips. These preliminary findings suggest that bionic reconstruction techniques can restore continuous neural control over multi-DOF prostheses to achieve agile locomotion over complex terrain.

**One-Sentence Summary:** A multi-DOF ankle-foot prosthesis under continuous neural control enables agile locomotion over complex terrain.

## INTRODUCTION

Walking is one of the most fundamental human abilities, emerging from the seamless integration of voluntary motor commands with reflexive spinal circuitry (*1–3*). Sensory information from muscles, tendons, joints, and the environment continuously informs these circuits, allowing the lower limbs to place the foot accurately, attenuate impact, generate propulsion, and recover from perturbations across diverse terrains (*1*, *4–7*). Unlike the dexterous movements of the upper limb, which are largely governed by volitional cortical control, locomotion depends heavily on reflex-driven modulation (*1*), enabling rapid adjustments crucial for balance and efficiency.

For people with standard-of-care limb amputation, replicating this adaptability remains a formidable challenge. Although powered prosthetic ankles and biomimetic controllers have advanced markedly in recent years (*8*, *9*), standard amputation procedures (*10*) sever substantial afferent and efferent neural pathways (*11*, *12*), limiting neural modulation of natural gait (*13–15*). To compensate, most neuroprosthetic systems employ intrinsic gait controllers—state machines or pattern-recognition algorithms that reproduce stored gait trajectories from onboard sensors (*13*, *16–20*). While these approaches can generate biomimetic mechanics on level ground, they typically restrict the user to high-level modulation of control parameters such as timing or gains, rather than granting direct, continuous neural control.

Direct stimulation of residual peripheral nerves has emerged as a complementary strategy for reintroducing afferent feedback after limb loss (*21*). Sophisticated stimulation paradigms can evoke biomimetic sensory signals and have enabled fine control of upper-extremity prostheses (*22*, *23*). However, locomotion is mediated largely by reflexive spinal pathways (*1–3*) with limited capacity for plastic reorganization compared to supraspinal networks (*24*). Effective restoration of lower-limb function may therefore require richer, more physiologically accurate afferent input. Recent studies have shown that sensory feedback can enhance the stability of passively or intrinsically controlled prosthetic legs (*25–29*), yet these devices remain dependent on internal control algorithms.

Recently, we demonstrated on persons with leg amputation that neuroprosthetic interfaces, electromyographic sensors, and mechatronic limbs can restore direct neural control over biomimetic and versatile gait, without relying on intrinsic gait controllers (*30*, *31*). Subjects with leg amputation were able to walk at peak speeds comparable to those of individuals without leg amputation and navigate real-world environments, including slopes, stairs, and obstructed pathways (*30*). The Agonist-Antagonist Myoneural Interface (AMI) (*30–33*), a surgical neuroprosthetic interface, enabled this advancement in neurally-controlled bionic gait. The AMI surgically links residual agonist-antagonist muscle pairs to replicate biological antagonistic muscle dynamics after amputation. This approach allows the use of native sensory organs (*11*, *12*) within residual muscles and tendons to provide neural afferents corresponding to free-space movements. While the AMI only emulates free-space movements, our previous works (*30*, *31*) have shown that the human nervous system can fine-tune residual motor control, achieving biomimetic gait through augmented residual afferents and continuous neural control authority over the bionic limb (Fig. 1A). As the level of residual afferent information (termed agonist-antagonist muscle afferents (*30*)) increases, individuals with leg amputation are better able to integrate with the bionic limb and achieve a higher degree of biomimetic gait (*30*, *31*).

**Fig. 1.**
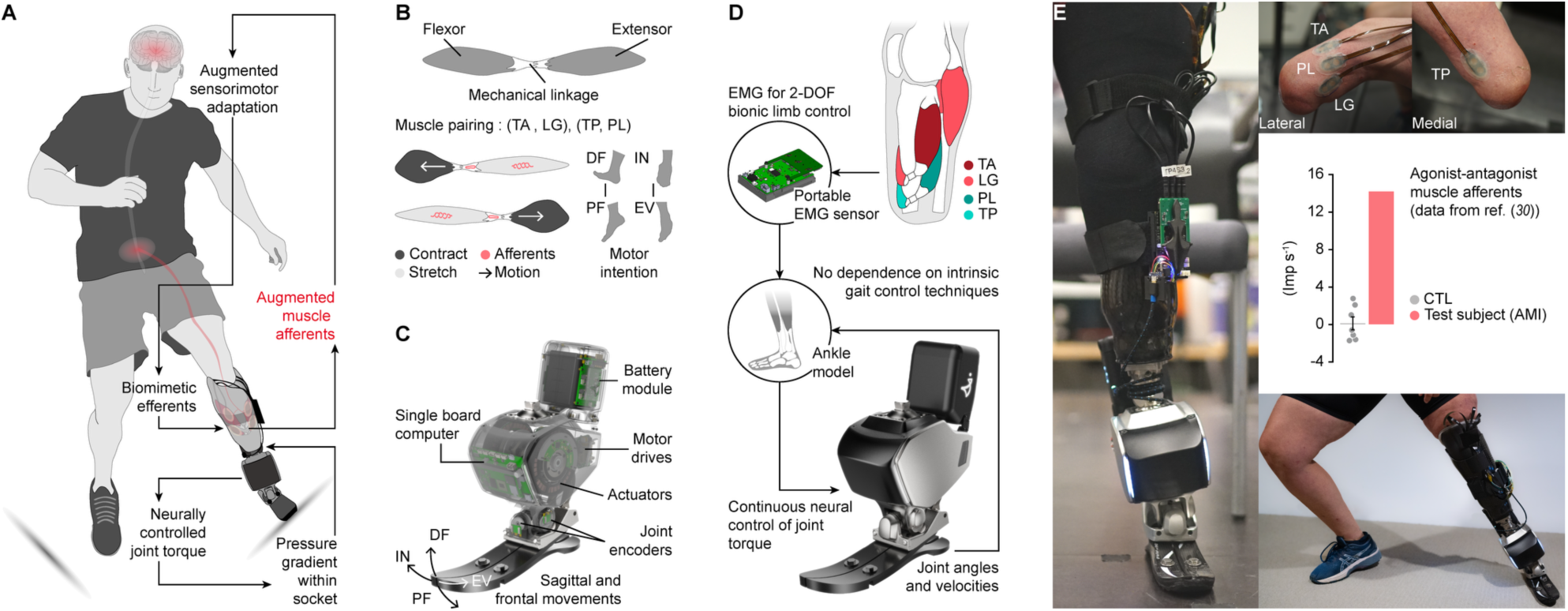
2-degree-of-freedom (DOF) bionic reconstruction. (**A**) Schematic of a 2-DOF bionic reconstruction for a robotic leg fully controlled by the human nervous system. The 2-DOF bionic reconstruction combines a robotic leg with a surgical neuroprosthetic interface designed to augment residual muscle afferents. This system facilitates sensorimotor adaptation to a biomimetic gait through continuous neural control and enhanced afferent feedback from residual muscles. Additional feedback may also be derived from mechanical information, such as pressure gradients within the prosthetic socket during ground contact. (**B**) Surgical neuroprosthetic interface: the Agonist-Antagonist Myoneural Interface (AMI). The interface preserves antagonistic muscle dynamics by mechanically linking agonist-antagonist muscle pairs during amputation. The AMI leverages native sensory organs in residual muscles to provide afferent feedback corresponding to muscle movements. Two AMIs were constructed using the tibialis anterior (TA), lateral gastrocnemius (LG), tibialis posterior (TP), and peroneus longus (PL) muscles for 2-DOF bionic control. DF, dorsiflexion; PF, plantar flexion; IN, inversion; EV, eversion. (**C**) The 2-DOF robotic ankle used in this study. (**D**) Continuous neural control paradigm. Electromyography (EMG) signals from the two AMIs (four muscles) were used to control the robotic ankle, granting full control authority to the human nervous system without reliance on intrinsic gait control techniques such as state machines or pattern recognition methods. (**E**) Implementation of the 2-DOF bionic reconstruction. The neuroprosthetic interface and robotic ankle were connected using flexible electrodes and a portable EMG sensor. Previous work (*30*) validated the augmentation of residual muscle afferents by the neuroprosthetic interface in the test subject. This reconstruction enables continuous neurally-driven control of the 2-DOF bionic movements.

These previous studies (*30*, *31*), however, focused on achieving the emulation of sagittal plane biomechanics under the full neural modulation of a single degree-of-freedom (DOF) leg prosthesis. While sagittal mechanics primarily govern gait energetics and terrain adaptation in various daily life settings, frontal joint mechanics play a critical role in maintaining stable gait across uneven terrains and during athletic activities (*34*, *35*). The frontal joint is particularly important for responding to external lateral perturbations, such as slips, which pose critical safety risks. A previous study (*32*) on frontal joint control was limited to free-space movements and walking at non-functional (semi-static) speeds, without demonstrating functional gait capabilities such as joint torque control or power generation. Consequently, the potential to restore direct neural control over multi-DOF bionic gait without relying on intrinsic gait control techniques remains an unresolved challenge.

In this preliminary study, we evaluate a neurally-modulated, 2-DOF ankle-foot prosthesis on one individual with a below-knee amputation, presenting case study results that show continuous neural control of both sagittal and frontal mechanics across level-ground walking, cross-slopes, dynamic obstacle maneuvers, and responses to unforeseen slips—all achieved without reliance on intrinsic gait control techniques.

## RESULTS

### Study design and participant

This study was part of a clinical trial (NCT03913273) investigating the potential of the Agonist-Antagonist Myoneural Interface (AMI) (*30–33*), a neuroprosthetic interface, to enhance multi-degree-of-freedom (DOF) neural control in individuals with below-knee AMI amputation. The AMI amputation surgically links residual agonist-antagonist muscle pairs to emulate intact muscle dynamics post-amputation. By replicating these dynamics, the AMI aims to leverage native sensory organs (*11*, *12*) embedded within the residual muscles and tendons, producing biomimetic afferents corresponding to free-space joint movements. In the below-knee AMI amputation, two AMIs are constructed using the tibialis anterior (TA) linked to the lateral gastrocnemius (LG), and the tibialis posterior (TP) linked to the peroneus longus (PL). These muscle pairs enable a 2-DOF bionic control (Fig. 1B). Our previous work (*30*) demonstrated that integration of these AMI constructs with surface electrodes, a control system, and a bionic ankle-foot prosthesis restored continuous neural control of biomimetic gait across various walking speeds in real-world environments, without relying on an intrinsic gait controller. This prior research focused on single-DOF sagittal ankle-foot mechanics, and thus the potential for simultaneous multi-DOF control remains unexplored. With a single study participant, this preliminary case study investigates the feasibility of biomimetic 2-DOF bionic control to restore both sagittal and frontal mechanics using a neurally-driven bionic ankle-foot prosthesis.

The study participant was a female in her 40s who had undergone a below-knee AMI amputation 5.1 years prior to the studies’ launch. She had previously participated in our study (*30*) on 1-DOF bionic gait with continuous neural control. Among individuals with amputations, the participant exhibited a high level of agonist-antagonist muscle afferents (*30*), a critical factor for facilitating augmented sensorimotor adaptation and enabling biomimetic bionic control. The study participant’s high afferent level made her a great candidate for exploring the potential of a 2-DOF bionic reconstruction. The test subject provided written informed consent prior to participation (IRB protocol 1812634918).

In this study, the gait trials were designed primarily to quantify 2-DOF bionic adaptation to uneven terrains. First, we conducted a level-ground walking study using a 10-meter walk test protocol to assess basic gait capabilities under continuous neural control, without any environmental perturbation. This validation confirmed that adding an additional DOF to the neuroprosthetic interface did not negatively impact the robotic limb’s stability under full neural control. Next, we tested 2-DOF bionic adaptation for inverted and everted cross-slope walking using a 15-degree slope block. Finally, the participant was asked to navigate a small obstacle course constructed from the four slope blocks, with the goal of hopping between blocks to clear the course. Both bionic mechanics and electromyography (EMG) used for bionic control were analyzed, along with video recordings, to assess biomimetic neural control of the 2-DOF bionic limb.

### 2-DOF bionic limb reconstruction

To test human-driven 2-DOF bionic gait, we developed and integrated a 2-DOF robotic ankle (Fig. 1C) with the neuroprosthetic interface, AMI (Fig. 1B) via skin mounted surface electromyographic (EMG) electrodes. The 2-DOF robotic ankle provides dorsiflexion (DF, 30 deg)–plantar flexion (PF, 50 deg) and inversion (IN, 35 deg)–eversion (EV, 35 deg) movements, with maximum torques of 240 Nm and 63 Nm, respectively. We extended the system architecture for 2-DOF bionic control based on our previous 1-DOF bionic reconstruction (*30*) (Fig. 1C). The same continuous neural controller from prior work was used for PF-DF torque control with an ankle model, and an additional neural controller was added for IN-EV joint stiffness control. Importantly, the controller design does not rely on intrinsic gait controllers, such as state machines or pattern recognition, which use biological gait knowledge to account for mechanics in gait phases, speeds, or terrain types. The neuroprosthetic interface (*30*, *32*, *33*), which augments muscle afferents, communicated with the robotic limb via flexible electrodes and a portable EMG sensor unit (*36*) (Fig. 1D). The bionic reconstruction followed the procedures from our previous study (*30*), but with four EMG channels used to record neural signals from the TA, LG, TP, and PL for 2-DOF control. Subject-specific neural decoding parameters were initially set based on settings from previous work (*30*), with further tuning performed based on feedback from the subject during the neurally-controlled 2-DOF walking practice session. Considering motor coordination among multiple muscles (*33*), the neural decoders can use neural inputs from all four muscles to determine motor intentions for each DOF control. However, for simplicity, the TA-LG muscle pair was used for DF-PF control, while the TP-PL muscle pair was used for IN-EV control. Once the subject reported feeling natural control over the bionic limb, the controller settings were fixed for the entire evaluation.

### Level-ground walking

We first tested level-ground walking at the subject’s preferred walking speed (1.32 ± 0.013 m s^-1^) along a 10-meter hallway (Fig. 2A). The subject achieved stable 2-DOF bionic walking under continuous neural control without requiring any external support, such as a handrail (Movie S1). For the sagittal plane, biomimetic dynamics (*4*) were observed, including a push-off during late stance followed by foot clearance in the swing phase (Fig. 2A). Additionally, peak PF power for propulsion (2.76 ± 0.23 W kg^-1^) and net work (0.267 ± 0.024 J kg^-1^) were normalized to typical physiological values reported in the literature (peak power: 3.0 W kg^-1^ (*37*), net work: 0.267 J kg^-1^ (*38*)) (Fig. 2B). In the frontal plane, a similar pattern of frontal adaptation to the biological ankle was observed (*39*), with inversion occurring during late stance. However, the peak inversion angle exceeded 20 degrees, which is notably higher than typical physiological values. To further understand how the 2-DOF bionic ankle was controlled by the human nervous system, we analyzed the EMG signals from the four AMI muscles (TA, LG, TP, and PL). The analysis revealed that the subject modulated the bionic ankle-foot prosthesis in a manner akin to the control of an intact limb during level-ground walking (*40*, *41*) (Fig. 2A, C). For sagittal neural control, push-off was achieved through a gradual increase in LG activity, while TA activation facilitated foot clearance and heel strike during early swing and early stance, respectively. For frontal neural control, the subject selectively coactivated TP and PL to increase joint stiffness for stability during leg support, while relaxing them during the swing phase to allow for a rapid transition and foot clearance. Our results underscore the potential for multi-DOF bionic reconstructive techniques to restore the neural control of multi-DOF biomimetic gait without relying on intrinsic gait controllers.

**Fig. 2.**
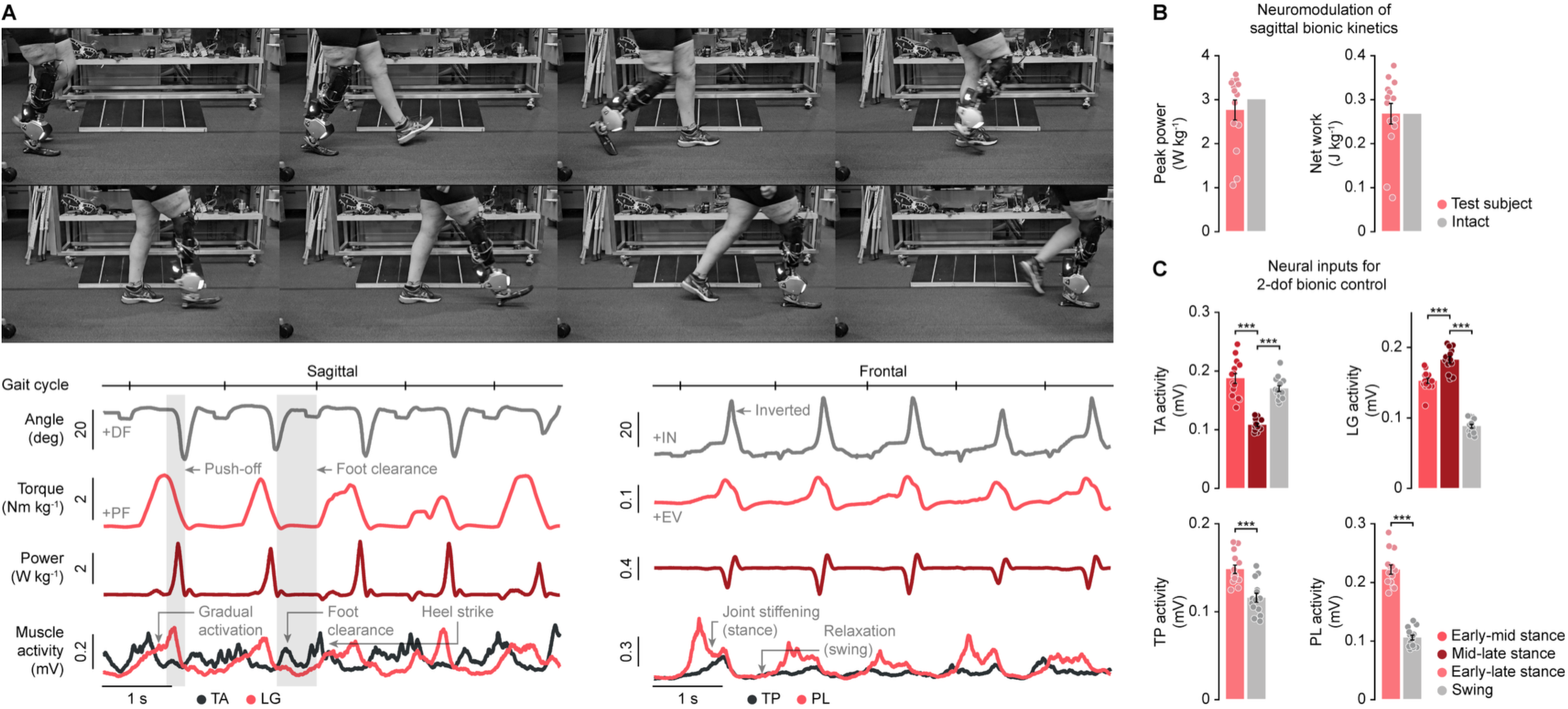
Level-ground walking with the 2-DOF bionic limb. (**A**) Chronophotography and 2-DOF bionic ankle mechanics during level-ground walking. (**B**) Normalized gait kinetics achieved through continuous neural control (bars, mean ± SEM; *N* = 14 steps). For comparisons, biological ankle kinetics from the literature are provided (*37*, *38*). (**C**) Biomimetic neural control for 2-DOF bionic gait (bars, mean ± SEM; *n* = 14 per phase; two-sided paired *t*-tests, \*\*\**P* < 0.00039). The TA is selectively activated during the early to mid-stance and swing phases, while the LG gradually increases its activation from early to mid-stance, peaks in late stance, and relaxes during swing. Both the TP and PL activate during the stance phase to stabilize the frontal joints, relaxing during the swing phase to enable a fast transition between stance and swing phases.

### Cross-slope adaptation

For a bionic limb to function effectively in real-world conditions, it must demonstrate the ability to respond and adapt to complex environmental perturbations (*8*). In previous work (*30*), a 1-DOF bionic reconstruction enabled individuals to adapt to stairs and slopes and to navigate obstructed pathways by performing obstacle crossings. In this study, we extended the investigation to test a 2-DOF bionic reconstruction for uneven terrain adaptation under full neural modulation. A 15-degree cross-slope block was used to simulate uneven terrain (Fig. 3A), with the slope oriented medially or laterally to create inverted or everted environmental perturbations on the bionic leg during walking in the same 10-meter hallway. The subject walked at a preferred speed and maintained this speed throughout the obstructed pathway, including during approach, adaptation, and recovery from the cross-slope. The subject demonstrated stable and biomimetic 2-DOF bionic cross-slope adaptation without requiring external support for either inverted or everted perturbations (Fig. 3A and Movie S2). The bionic limb adapted to the cross-slope by inverting or everting the joint, depending on the direction of the perturbation, while simultaneously increasing joint torque in the opposite direction to provide stability against the environmental perturbations (Fig. 3B, C). EMG analysis revealed that this adaptation was achieved by selectively increasing the coactivation of the TP and PL muscles during the stance phase, resulting in greater joint stiffness on the cross-slope block compared to unobstructed steps (Fig. 3D). Despite the increased coactivation of the TP and PL muscles during the stance phase for cross-slope adaptation, the subject was able to relax the muscles during the swing phase, facilitating a fast transition for foot clearance and positioning. Additionally, the subject exhibited distinct neural control patterns for inverted and everted cross-slope adaptation (Fig. 3E). For inverted perturbations, which required increased eversion torque, the coactivation was dominated by heightened activation of the PL, a biological evertor muscle. Conversely, for everted perturbations, which demanded increased inversion torque, coactivation was achieved with less pronounced PL activation. These findings indicate that the subject did not merely increase the overall level of muscle coactivation but instead employed biomimetic motor strategies tailored to each specific environmental perturbation, enabling effective bionic responses.

**Fig. 3.**
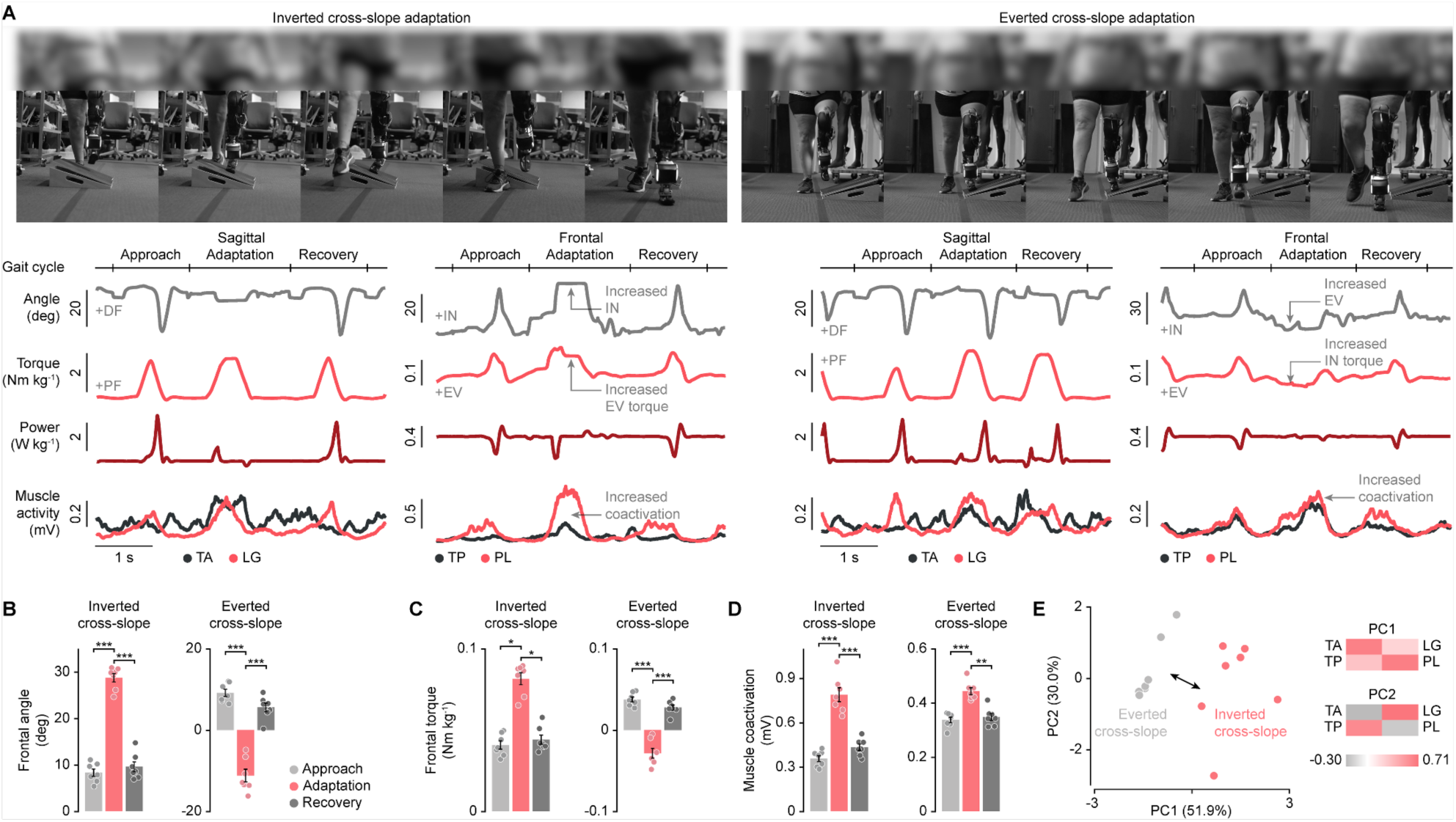
Cross-slope adaptation with the 2-DOF bionic limb. (**A**) Chronophotography and 2-DOF bionic ankle mechanics during cross-slope adaptation. (**B**) Adaptative bionic kinematics and (**C**) kinetics (bars, mean ± SEM; *N* = 7 per phase; inverted cross-slope frontal torque comparisons: Wilcoxon tests, others: two-sided paired *t*-tests, \**P* = 0.016, \*\*\**P* < 0.00015). For an inverted slope, the frontal angle is inverted to adapt to the slope, accompanied by increased eversion torque to stabilize the joint. Conversely, for an everted slope, the frontal angle is everted, followed by increased inversion torque to ensure joint stabilization. (**D**) Neural control of adaptive mechanics during the stance phase (bars, mean ± SEM; *N* = 7 per phase; two-sided paired *t*-tests, \*\**P* = 0.0014, \*\*\**P* < 0.00084). The subject demonstrated selectively increased muscle coactivation levels during side-slope adaptation compared to steps before and after the slopes. (**E**) Principal component analysis (PCA) of the four muscle EMG signals reveals distinct neural control strategies for each type of obstructed pathway. PC1 and PC2, principal component 1 and 2.

### Dynamic obstacle course maneuver: Bionic Angled Dash

Based on the promising results of the bionic reconstruction, we expanded our investigation to more athletic scenarios. A small obstacle course, *Bionic Angled Dash*, was constructed using four 15-degree cross-slope blocks (Fig. 4A), inspired by the Angled Dash from American Ninja challenge (*42*). The blocks were oriented medially, with two blocks for each limb side. The course was designed to require the subject to complete two full gait cycles for each limb side, stepping on the blocks using either the intact or bionic limb independently. The subject was instructed to hop between the blocks without touching the ground, while no specific guidance on bionic control strategies was provided. The subject successfully completed the obstacle course, demonstrating sequential single-leg balancing and precise foot positioning with the bionic limb on the cross-slope blocks (Movie S3). For sagittal plane mechanics, the subject controlled their bionic limb similarly to their intact limb when maneuvering through the course. The subject positioned the bionic limb by performing PF during the swing phase before stepping onto the block (toe landing), mirroring the motion of their intact limb (Fig. 4A-C). This adaptive foot positioning was facilitated by a selective increase in LG activity during the swing phase prior to block contact (Fig. 4D). Peak PF torque gradually decreased as the subject progressed through the course (Fig. 4B, C). However, both sagittal plane kinematics and kinetics returned to levels observed during unobstructed gait immediately after the course (Fig. 4B, C). For frontal plane mechanics, the subject modulated the bionic limb in a manner consistent with the single-cross-slope adaptation trial. For the first block, the ankle was everted while the inversion joint torque was increased (Fig. 4B, C). However, on the second block, the frontal kinematics shifted closer to a neutral position, showing less inversion compared to the first block (Fig. 4B, C). This adjustment was likely due to a change in thigh orientation, as the first block required a ground-to-block transition, while the second involved a block-to-block transition (Movie S3). This alignment reduced the demand for ankle eversion during the second block adaptation. Muscle coactivation notably increased during the two-block adaptation phase and slightly during the recovery step (Fig. 4B, D). This increased coactivation likely served to increase joint stiffness for single-leg support on the blocks and to manage the impact of transitioning from the block to the ground. Both frontal plane kinematics and kinetics largely returned to baseline levels of unobstructed walking immediately after the course (Fig. 4B, C). Our results suggest that bionic reconstruction may enable neurally-driven bionic limbs to extend beyond regular daily activities, such as walking on level ground or stairs, and potentially provide support during athletic activities.

**Fig. 4.**
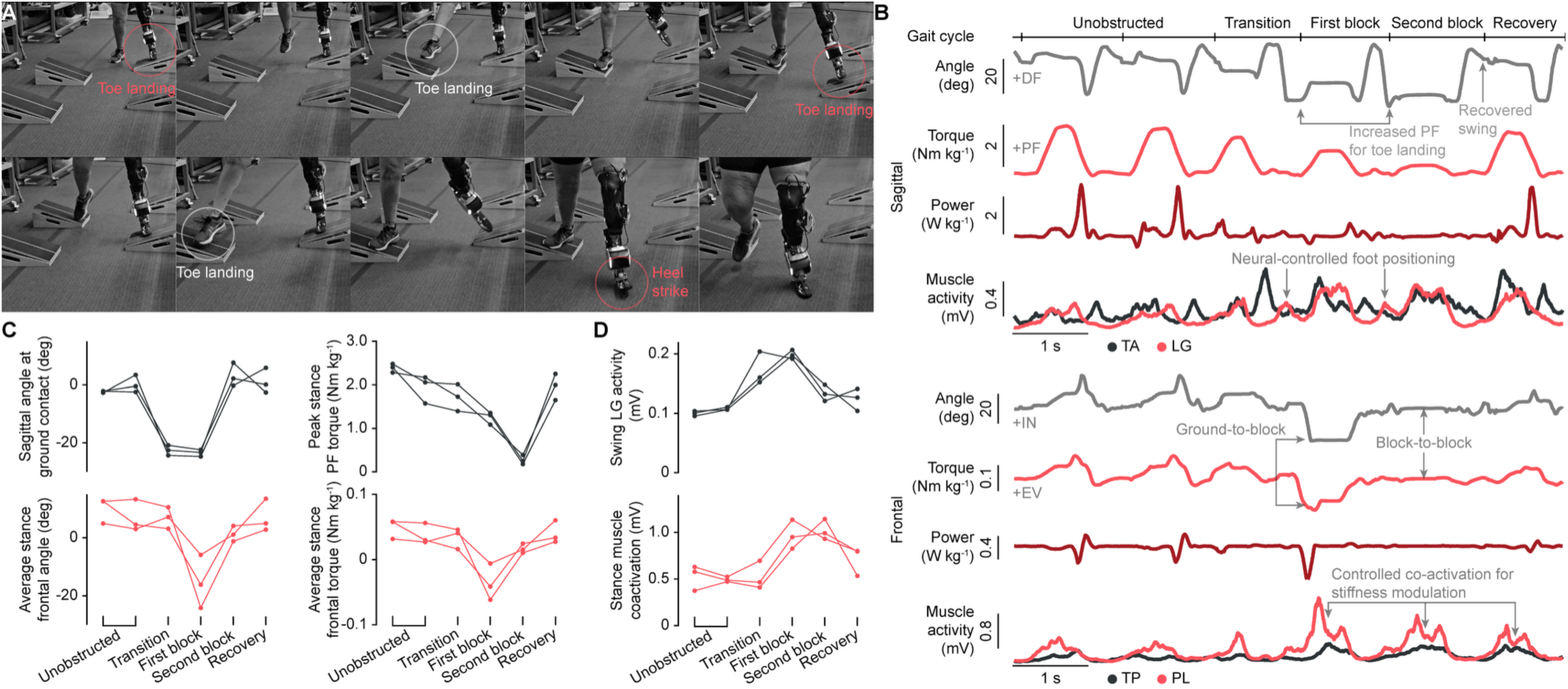
Obstacle course maneuver with the 2-DOF bionic limb. (**A**) Chronophotography and (**B**) 2-DOF bionic ankle mechanics during an obstacle course maneuver. The course consisted of four cross-slope blocks, with two blocks for each limb side, positioned in an everted orientation. Both the intact and bionic limbs exhibited toe landing on each block, followed by a heel strike of the bionic limb to facilitate immediate recovery after clearing the obstacle. (**C**) Bionic mechanics and (**D**) neural control during the maneuver (*N* = 3 trials). Swing phase adaptations primarily at the transition and first block steps, while stance phase adaptations occurred primarily at the first and second block steps. The neurally-controlled mechanics recovered to regular level-ground walking immediately following the maneuver.

### Bionic response to an unforeseen slip

During the Bionic Angled Dash trial, an unexpected incident occurred when the second block for the bionic limb slipped laterally due to suboptimal fixation (Fig. 5A, Movie S4). The slip happened while only the bionic limb was in contact with the block, and the intact limb was in mid-air, preparing to step onto the next block. Remarkably, the subject was able to respond to this slip, maintain balance, and quickly reposition both the bionic and intact limbs. Initially, the subject attempted to stabilize on the slipping block by rapidly increasing PF torque and stiffening the joint through overall muscle coactivation (Fig. 5B). This provided enough time for the subject to successfully land the intact limb on the next block without falling. As the block continued to slip and maintaining the bionic limb on it became increasingly difficult, the subject quickly relaxed the bionic ankle, repositioned the limb, and ramped up muscle activation again. This adjustment allowed the bionic limb to provide stable support, enabling the intact limb to follow and the subject to regain a stable standing position within her comfort zone. Although this observation stemmed from an unexpected incident, it demonstrates the capability of the bionic reconstruction on this subject to reflexively respond to unforeseen dynamic perturbations that might have otherwise led to a fall.

**Fig. 5.**
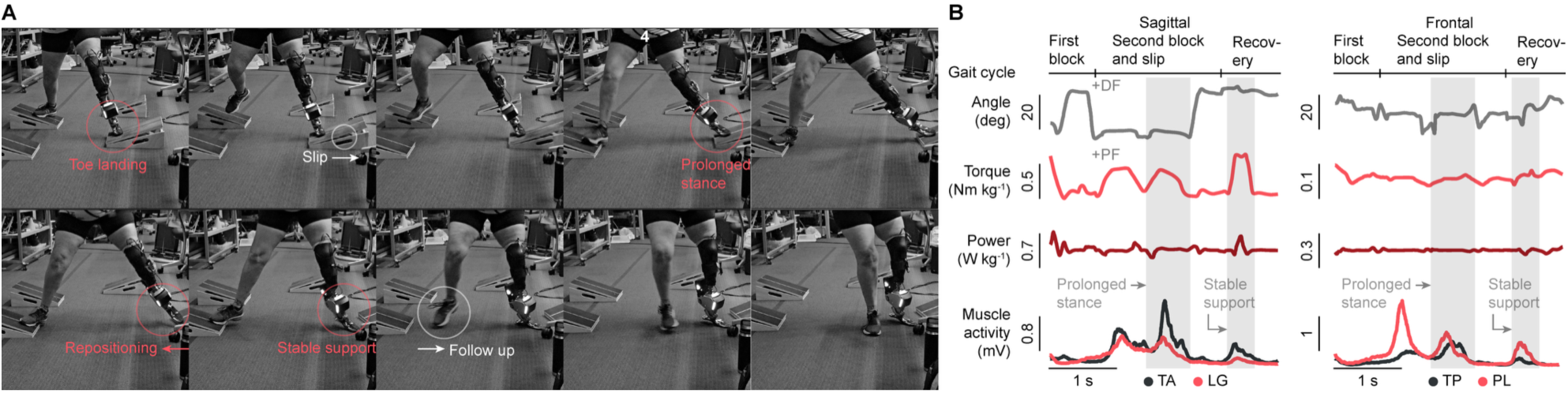
Bionic response to an unexpected obstacle block slip. (**A**) Chronophotography and (**B**) 2-DOF bionic ankle mechanics during an unexpected obstacle block slip. The neurally-controlled bionic limb responded effectively to the slip by repositioning and stabilizing the limb, thereby preventing a potential fall.

## DISCUSSION

In this work, we demonstrate that a 2-DOF bionic reconstruction can restore direct human-driven control of bionic legs capable of not only level-ground walking and basic urban slopes and stairs, but also cross-slope adaptation, dynamic obstacle maneuvering, and effective responsiveness to unforeseen slips. To the best of our knowledge, even with advanced intrinsic control systems (*8*, *9*), the ability to perform dynamic obstacle maneuvers and respond to slips, as achieved in this study, remains unprecedented in the existing literature. Building on our previous findings with 1-DOF bionic leg reconstruction (*30*), this work provides preliminary evidence that surgically reconstructed agonist-antagonist muscles, which leverage native muscle-tendon sensory organs to provide afferents (*11*, *12*), are effective for enabling 2-DOF bionic legs with continuous neural control.

In previous work (*30*, *31*), we demonstrated that humans could refine their residual motor control to integrate with a bionic limb through the amplification of native residual afferent signaling combined with continuous neural control authority for a 1-DOF bionic control. In this study, we extend the same bionic reconstruction framework to enable a 2-DOF bionic control, demonstrating its effectiveness in restoring human control authority over a bionic limb. However, applying this framework to different formats of 2-DOF bionic control, such as bionic ankle-knee systems, as opposed to the sagittal-frontal bionic ankle-foot control described here, may present unique challenges. For instance, controlling sagittal-frontal bionic ankle-foot mechanics demands higher bandwidth control compared to more proximal joints (*4*, *5*), where the coordination of bionic ankle-knee mechanics requires precise temporal alignment, such as ankle-powered plantarflexion during late stance and its influence on early swing-knee kinematics (*43*). Additionally, the principles of bionic reconstruction that leverage human sensorimotor adaptation through amplified residual afferent signaling may vary depending on the specific neural deficits associated with different levels of amputation. Indeed, future research exploring the application of various bionic reconstruction frameworks to diverse joint configurations may provide deeper insights into how the human nervous system integrates with bionic systems, potentially broadening the scope of functional restoration.

Previously, we demonstrated that increasing residual afferent information (termed agonist-antagonist muscle afferents (*30*)) enhanced integration with a bionic limb, enabling more biomimetic gait, greater neural control bandwidth, and prosthetic embodiment (*30*, *31*). The study subject in this work was among the participants with high afferent levels from our earlier study (*30*). In this preliminary study, the participant’s high residual afferent level provided favorable conditions to investigate the potential for a 2-DOF bionic reconstruction. Building on the principles of our 1-DOF bionic reconstruction research (*30*, *31*), we anticipate that individuals with higher residual afferents would achieve biomimetic capabilities and control bandwidth for multi-DOF bionic limbs beyond those previously reported. In contrast, more limited bionic control is expected in individuals with lower residual afferents. Although the findings of this exploratory study are limited by its case study design, future research involving larger populations with varying afferent levels could further elucidate the role of residual afferent information in achieving effective multi-DOF bionic reconstruction.

In this study, the 2-DOF neural controller utilized two AMI agonist-antagonist muscle pairs as independent control inputs for each DOF based on their primary functions. Specifically, the EMG signals from the TA-LG muscle pair were used for DF-PF control, while those from the TP-PL muscle pair were used for IN-EV control. This simplified approach enabled a quick calibration process, allowing gait testing to be performed efficiently within clinical constraints, such as minimizing patient fatigue. Meanwhile, as residual afferents increase, residual motor control in individuals with amputation is known to approximate the motor control observed in individuals without amputation during free-space tasks, where all four muscles contribute to both DOFs through muscle synergies (*33*). Consequently, neural decoding performance could potentially be improved by designing a neural controller that incorporates neural signals from all four muscles to decode each DOF. The neural controller in this study was initially set using auto-tuned parameters derived from maximum PF and DF measurements during standing from the previous study (*30*) and further refined based on subject feedback during the usage of the 2-DOF bionic leg in this study. Additionally, a commonly used bi-linear relationship between EMG signals and decoding variables, target joint angle and impedance regulation level, was employed (*30*, *44*, *45*). While this calibration process and simplified neural decoding approach were sufficient for the initial validation of the 2-DOF bionic reconstruction, advanced methods, such as human-in-the-loop calibration combined with non-linear decoding techniques (*46*), may further enhance bionic controllability. In such future implementations, utilizing sensing platforms that minimize or eliminate crosstalk between muscle control signals, such as magnetomicrometry (*47*), could enhance multi-DOF bionic reconstruction and control.

A previous study (*32*) on 2-DOF bionic control was conducted only at non-functional (semi-static) walking speeds and failed to demonstrate key functional gait mechanics, such as joint torque modulation and power generation. Consequently, restoring continuous neural control of a 2-DOF biomimetic bionic gait has remained an unresolved challenge. The 2-DOF bionic control paradigm in this study enables continuous neural control of DF-PF joint torque, as well as IN-EV joint stiffness. Given the potential limitations of the simplified decoding model, this design was primarily chosen to address safety concerns in this initial exploratory investigation. Despite this conservative design, the subject demonstrated the ability to achieve biomimetic level-ground walking, dynamic obstacle maneuvers, and responses to unforeseen slips without relying on an intrinsic gait controller, a notable outcome given the simplified decoding framework. During single cross-slope block adaptation trials, the subject demonstrated distinct motor outputs to adapt to inverted and everted blocks, corresponding to an increase in eversion and inversion joint torque, respectively. This observation suggests the capability to generate neural control inputs for more direct control of the IN-EV joint through a torque control rather than the stiffness modulation approach pursued in the present study. Future work could focus on expanding the decoding methodology to include IN-EV joint torque modulation and exploring its applications beyond the locomotion tasks demonstrated in this study.

During the Bionic Angled Dash challenge, the subject naturally adopted symmetric bionic control with the intact limb for toe landing, despite receiving no specific instructions to do so. Given the uniqueness of the tested tasks, we cannot determine whether the observed toe landing transitions between blocks represent general motor strategies for the intact population nor other individuals with limb amputation who possess a high level of bionic reconstruction. However, it is noteworthy that the subject was able to project the bionic limb in kinematics similar to an intact limb. Furthermore, to our knowledge, the capability of a bionic limb to perform such dynamic obstacle maneuvers has not been reported in existing literature, even with intrinsic gait control. The bionic locomotion demonstrated in this study establishes a foundation for testing direct human-driven bionic capabilities beyond regular daily activities, perhaps extending future testing to athletic pursuits.

During one of the Bionic Angled Dash trials, a cross-slope block slipped laterally due to suboptimal obstacle course settings. This occurred at a critical moment when the bionic limb was providing single leg support on the block, while the intact limb was mid-air transitioning between blocks. Despite this unexpected event, the subject rapidly adjusted the bionic limb mechanics, repositioned, and provided stable support for the intact limb, successfully recovering balance.

This incident highlights a critical capability of the bionic reconstruction in real-world applications, namely the ability to actively respond to critical events and recover to a safe state. Future studies that systematically investigate neurally-controlled bionic responses to such dynamic events could offer valuable insights into the bionic reconstruction framework required for safe and reliable locomotion.

The realization of direct, neurally-driven prosthetic limbs capable of biomimetic and versatile gait has long been a goal in the field of bionics. Recently, bionic reconstruction techniques have emerged that enable individuals with limb loss to regain direct control over their locomotion. This work provides preliminary evidence that multi-DOF bionic locomotion driven by human intent is achievable. Furthermore, the study results suggest that such systems can support not only routine daily activities, but also athletic tasks necessitating critical safety responses, all without dependence on intrinsic gait controllers.

## MATERIALS AND METHODS

### Experimental design

This study is part of a clinical trial (NCT03913273) investigating the potential of the Agonist-Antagonist Myoneural Interface (AMI) (*30–33*), a neuroprosthetic interface, to enhance multi-degree-of-freedom (DOF) neural control in individuals with below-knee AMI amputations. The AMI amputation procedure reconstructs agonist-antagonist muscle dynamics by surgically linking major residual muscle pairs, replicating biological mechanoneural transduction (*11*, *12*). This allows leveraging native sensory organs in the residual muscle-tendon complex to provide afferent proprioceptive signaling for free-space joint movements. Building on prior findings of 1-DOF continuous neural control of biomimetic gait (*30*), this exploratory study investigates whether similar neurally-controlled biomimetic gait can emerge for a 2-DOF bionic reconstruction. Previous research (*30*) demonstrated that the 1-DOF bionic reconstruction comprising the AMI amputation procedure with a bionic limb restores neurally-controlled biomimetic gait without relying on an intrinsic gait control. Among varying levels of residual afferent information (termed agonist-antagonist muscle afferents (*30*)), individuals with higher afferent levels achieved more effective integration with the 1-DOF bionic limb, demonstrating greater biomimetic gait and control bandwidth. This study aimed to evaluate the potential of a 2-DOF bionic reconstruction by: (1) developing an autonomous 2-DOF bionic ankle enabling control of sagittal and frontal ankle and subtalar joint mechanics; (2) integrating the bionic system on an individual with a neuroprosthetic interface, namely the AMI; (3) assessing 2-DOF bionic gait with continuous neural control during level-ground walking, a single cross-slope block adaptation, and dynamic obstacle course maneuver; and (4) performing biomechanical and EMG analyses to quantify the degree of biomimetic gait and explore underlying neural control strategies. Video recordings of the bionic gait trials were analyzed to compare movements of the intact limb with those of the bionic limb. A single participant, previously involved in the 1-DOF bionic gait study (*30*), was recruited. This participant’s high afferent level made her an ideal candidate for exploring the potential of a 2-DOF bionic reconstruction. Written informed consent was obtained prior to participation (IRB protocol 1812634918). No control group was included; comparisons were made with typical motor control reported in the literature and the participant’s intact limb, as observed in video recordings.

### 2-DOF bionic reconstruction

We developed a 2-DOF bionic leg consisting of a powered prosthetic ankle integrated with the subject’s neuroprosthetic interface via a portable EMG sensor system (weight: 57 g) and flexible bipolar surface electrodes (width: 18 mm, length: 60 cm) (*36*). The powered prosthetic ankle-foot prosthesis is capable of generating active joint torque up to 240 Nm for the sagittal (DF-PF) DOF and 63 Nm for the frontal (IN-EV) DOF, using two brushless electric motors (U8 Lite KV100, T-motor) with a 39:1 gear ratio. Low-level torque control was implemented by back-calculating target force values from the target torque using the moment arm and transmission ratio, which were then achieved through proportional-integral control based on current feedback. The prosthesis provides a range of motion (ROM) of 30 deg DF, 50 deg PF, and 35 deg IN and EV. The prosthesis measures 200 mm in height, 104 mm in width, and weighs 3.4 kg, including a 0.3 kg battery module capable of powering the system for approximately 2 hours of continuous usage per charge. The bionic reconstructive process followed the procedures outlined in our previous work (*30*). Briefly, to enable robust EMG recording within the socket, flexible bipolar electrodes were used with four channels to record signals from the TA, LG, TP, and PL muscles for 2-DOF bionic control, expanding upon earlier work (*30*) which used only the TA and LG muscles. Each electrode was affixed to its target muscle using double-sided tape, electrode gel (SPECTRA 360, Parker Labs Inc.), and a hydrocolloid gel patch (Pnrskter blister bandages). The electrodes and their lead wires were routed through a liner-liner prosthetic sock and prosthetic liner to prevent skin irritation or damage from friction. After electrode placement, the subject donned their prosthetic socket, and the 2-DOF powered ankle-foot prosthesis was attached. The electrodes were then connected to the EMG sensor, which was secured to the side of the socket and linked to the powered prosthesis. EMG signals were processed in real time following the same methodology as in the previous study (*30*) for 2-DOF bionic control. Raw EMG signals were recorded at 2 kHz and band-passed using a finite impulse response (FIR) filter (stop-band: 0-60 Hz, >360 Hz; pass-band: 90-330 Hz; stop-band attenuation: 85 dB; order: 198) and a cumulative histogram filter (*48*) to ensure robust readings within the liner-socket system. The root-mean-square (RMS) of rectified EMG signals (200 ms window size) was normalized using minimum and maximum values to compute the EMG envelope. Muscle activity was calculated from the EMG envelope using bilinear muscle activation dynamics (time constants: *t*_act_ = 10 ms, *t*_deact_ = 50 ms) (*49*).

### 2-DOF bionic control design

To enable continuous neural control of sagittal and frontal ankle mechanics, we expanded the 1-DOF continuous neural controller from prior work (*30*). The previous controller was developed based on the impedance control paradigm (*44*, *45*), incorporating joint position- and velocity-dependent torque characteristics of a biological ankle joint (*50*, *51*). The same controller was adopted for sagittal joint torque (*τ*_s_), offering continuous neural control of bionic DF-PF as

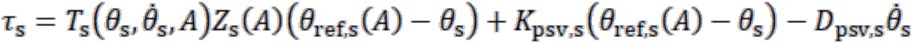

where 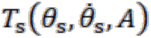 is the upper bound maximum torque for a given prosthetic sagittal angle (*θ*_s_) and angular velocity 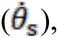 derived from biological joint torque data reported in the literature. It coordinates TA and LG muscle activity (*A*) to account for the different upper bound maximum torques for DF and PF, depending on the direction of actuation. The actuation direction is determined by the sign of difference between the neural-controlled target sagittal joint position (*θ*_ref,s_(*A*)) and *θ*_s_.*Z*_s_(*A*)indicates the impedance regulation level (value between 0 and 1), while and *K*_psv,s_ and *D*_psv,s_ represent passive sagittal joint stiffness and damping, respectively. The first term calculates the neural-controlled DF-PF joint torque by computing the maximum active joint torque for a given prosthetic angle and velocity, which is then scaled based on the motor intention, including the direction and intensity of the joint movement. For detailed formalization, refer to (*30*). For simplicity, *Z*_s_(*A*) and *θ*_ref,s_(*A*) are computed using the weighted sum and difference of TA-LG activity, respectively. This design enables direct, continuous neural control of DF-PF joint torque while mimicking the angle- and velocity-dependent properties of biological joint. The frontal joint torque (*τ*_f_) was neurally-controlled through joint stiffness modulation as

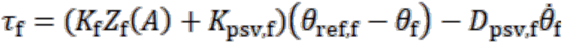

where *K*_f_ is the frontal active joint stiffness scaled by the frontal impedance regulation level (*Z*_f_(*A*)).*θ*_ref,f_ and *θ*_f_ represent the frontal target joint position with fixed at the neutral position and prosthetic position, respectively. *K*_psv,f_ and *D*_psv,f_ denote passive frontal joint stiffness and damping, respectively. Similar to the sagittal joint, *Z*_f_(*A*) was computed using the weighted difference of TP-PL activity. Each decoding parameter set for *Z*_s_(*A*), *Z*_f_(*A*), and *θ*_ref,s_(*A*) was initially determined based on the prior study (*30*) and subsequently fine-tuned based on user feedback and observations during practice runs before data collection.

### Level-ground walking experiment

To ensure safety, the participant first adapted to the 2-DOF bionic limb on a 3.6-meter walkway with handrails. The participant was advised to use the handrails whenever needed to assist with balance while walking at a self-selected speed. Additionally, the participant was asked to control the bionic gait in a way that felt natural and to provide feedback to the research team for fine-tuning the controller. Once the participant demonstrated steady, stable walking, testing progressed to a 10-meter open hallway without handrails. Given her prior experience with continuous neural control of a 1-DOF bionic limb (*30*), the participant expressed confidence in walking with the 2-DOF bionic limb on the 10-meter walkway without requiring long-duration practice sessions, such as those conducted in the prior study (∼6 hours) (*30*). However, because 2-DOF bionic limbs require simultaneous control of sagittal and frontal mechanics for steady walking, the risk of falling is inherently higher compared to the 1-DOF bionic walking tests. Consequently, in this exploratory case study, the participant was advised to walk at a preferred speed without a specific target walking speed.

### Bionic gait data collection and analysis

Bionic joint kinematics, torque, and EMG data were recorded using onboard sensors and transmitted to a laptop via telemetry at 500 Hz. A total of 14 gait cycles were collected for level-ground walking evaluation, with each gait cycle segmented using a sagittal torque threshold of 5 Nm. Biomimetic kinematic features, including push-off during the stance phase and foot clearance during the swing phase, were assessed. Bionic kinetics were evaluated by calculating peak positive power and net work from recorded kinematics and torque data, and these values were compared with typical biological data reported in the literature (*37*, *38*). To analyze underlying neural control strategies for 2-DOF bionic gait, sagittal neural control was assessed using TA and LG EMG data. The TA muscle is primarily responsible for controlled PF following heel strike and active foot clearance during early swing, while the LG muscle gradually increases activity throughout the stance phase for push-off and becomes silent during the swing phase to facilitate a rapid transition for foot clearance (*40*, *41*). Accordingly, average TA and LG EMG values were computed for three gait phases: early-mid stance (0–50% stance), mid-late stance (50–100% stance), and swing phases. Biological ankles increase overall joint stiffness during the stance phase compared to the swing phase (*40*, *41*). Therefore, frontal neural control was assessed by comparing TP and PL activation levels during early-late stance and swing phases. Walking speed was estimated based on travel time and distance.

### Cross-slope adaptation assessment

After the participant demonstrated the ability to perform stable 2-DOF bionic walking, we tested the potential of the 2-DOF bionic reconstruction to adapt to uneven terrain using a single 15-degree cross-slope block. Cross-slope adaptation trials were conducted in the same 10-meter open hallway. The participant was instructed to walk at their maximum self-selected speed while traversing the cross-slope block, which was positioned at the midpoint of the hallway. Both inverted and everted cross-slope scenarios were tested by placing the slope medially and laterally, respectively. Bionic ankle behavior was assessed during approach, adaptation, and recovery steps. A total of 7 adaptations for each cross-slope type were collected for evaluation. Frontal adaptive bionic kinematics and kinetics were analyzed by comparing the average joint angle and torque during the stance phase. To determine whether adaptive bionic mechanics resulted from passive impedance or active neuromodulation, the sum of TP and PL muscle activities during the stance phase were computed for each step. Additionally, to evaluate whether the participant employed general muscle coactivation or selectively adjusted muscle activity to adapt to different terrain types, principal component analysis (PCA) was applied to the average EMG data of the four AMI muscles during the stance phase. Each muscle’s EMG dataset was normalized using its mean and standard deviation before applying PCA. The PC1 and PC2 scores were used to represent the neural control strategies for inverted and everted cross-slope adaptations.

### Dynamic obstacle maneuver assessment

To explore the 2-DOF bionic reconstruction in more athletic scenarios, we constructed a small obstacle course, the *Bionic Angled Dash*, inspired by the Angled Dash from the American Ninja challenge (*42*). The *Bionic Angled Dash* consisted of four 15-degree cross-slope blocks oriented medially to create everted obstacles. Two blocks were placed on each side to allow testing two full gait cycles per limb, including transitions between blocks. The distances between blocks were set to approximately match the participant’s stride length. The participant was instructed to traverse the obstacle course without touching the ground between blocks, but was not provided specific instructions on bionic control strategies. After two practice trials, four obstacle maneuver trials were recorded. To assess how the participant controlled the bionic limb compared to their biological ankle-foot complex, the obstacle maneuvers were video-recorded. Among the four trials, one included an unexpected slope slip, which is analyzed in the next section. For the remaining three trials, six sequential steps were analyzed: two steps on an unobstructed pathway, a transition step from level ground to the first block, two steps on the cross-slope blocks, and a recovery step from the block to the ground. Although no explicit instructions for bionic control were given, the participant naturally mirrored biological ankle foot positioning by increasing PF with the bionic limb during the swing phase of the obstacle maneuver. To evaluate neurally-controlled bionic limb positioning, the sagittal bionic angle at ground contact and the average LG activity level during the swing phase were computed. Additionally, peak bionic PF torque was calculated to assess the ability to modulate propulsion during the obstacle maneuver and in steps before and after encountering the obstacle. For frontal mechanics, as in the single cross-slope adaptation trials, the average frontal angle, torque, and TP and PL coactivation levels during the stance phase were computed.

### Analysis of bionic response to an unforeseen slip

During one of the obstacle maneuver trials, a cross-slope block serving as the second block for the bionic limb step unexpectedly slipped laterally due to sub-optimal fixation. This unexpected event allowed for an evaluation of the bionic response based on changes in joint kinematics, torque, and EMG activity of the four AMI muscles at the moment of the block slip. Additionally, the participant’s overall locomotive response to the slip was assessed using video recordings.

### Statistics

All bionic gait and EMG data were reported as mean ± standard error of the mean (SEM). Data normality was verified by the Shapiro-Wilk test (significance level *α* = 0.05). For normally distributed data, two-sided paired *t* tests were used for within stride comparisons. For data that violated normality, Wilcoxon tests were applied. Statistical analyses were conducted using MATLAB 2023b (Mathworks, USA).

## Data Availability

All data are available in the main text or the supplementary materials.

## Supplementary Materials

Materials and Methods

Movies S1 to S4

## Acknowledgments

We thank M. Gonzalez for clinical support during the bionic gait data collection.

## Author contributions

T.H., H.S., and H.M.H. conceived the study concept. T.H., H.S., C.H., and H.M.H. contributed to experimental methods and neuroprosthetic control design. T.H designed the 2-DOF robotic ankle. T.H., H.S., C.H., D.V.L., S.H.Y., J.Q., and T.S. implemented the neuroprosthetic system. T.H., H.S., and C.H. conducted bionic gait studies. H.S. analyzed experimental data. M.J.C. conducted the AMI amputation procedure. H.M.H. performed study management and oversaw project funding. T.H., and H.S. prepared figures and wrote the original draft. T.H., H.S., and H.M.H. edited the manuscript. All authors revised and approved the final manuscript.

## Data and materials availability

All data are available in the main text or the supplementary materials.

